# Daily viral kinetics and innate and adaptive immune responses assessment in COVID-19: a case series

**DOI:** 10.1101/2020.07.02.20143271

**Authors:** Pauline Vetter, Christiane Eberhardt, Benjamin Meyer, Paola Andrea Martinez Murillo, Giulia Torriani, Fiona Pigny, Sylvain Lemeille, Samuel Cordey, Florian Laubscher, Diem-Lan Vu, Adrien Calame, Manuel Schibler, Frederique Jacquerioz, Géraldine Blanchard Rohner, Claire-Anne Siegrist, Laurent Kaiser, Arnaud M Didierlaurent, Isabella Eckerle

## Abstract

**Background:** Viral shedding patterns and its correlation with the immune responses of mildly symptomatic COVID-19 patients are still poorly characterized.

**Methods:** We enrolled the first five COVID-19 patients quarantined in our institution; none received immunomodulatory treatment. We monitored shedding of viral RNA and infectious virus by RT-PCR and cell culture from the upper respiratory tract, and characterized the kinetics of systemic innate and adaptive immune responses.

**Results:** Despite mild clinical disease, high viral loads and shedding of infectious virus were observed from the respiratory tract, with isolation of infectious virus and prolonged positivity by PCR up to day 7 and 19 post onset of symptoms, respectively. Robust innate responses characterized by an increase in activated CD14+CD16+ monocytes and cytokine responses were observed as early as 2 days after symptoms onset. Cellular and humoral SARS-CoV-2 specific adaptive responses were detectable in all patients.

**Conclusion:** Infectious virus shedding was limited to the first week of symptom onset in mild cases. A strong innate response, characterized by the mobilization of activated monocytes during the first days of infection, as well as SARS-CoV-2 specific antibodies were detectable, even in patients with mild disease.

**Summary:** We describe viral and immune profiles of the first five SARS-CoV-2 patients in our institution, showing high viral loads and infectious viral shedding in early acute disease. Mild patients mount an innate response sufficient for viral control and specific immunity.

## Introduction

As of 18th June 2020, more than 8 million cases of COVID-19 have been reported worldwide, with 450,000 deaths [2]. While the first reports mainly described patients presenting with severe pneumonia [3], the spectrum of disease is wide, with more than 80% of infected individuals displaying moderate, mild or even no symptoms [4, 5].

Studies to date have focused on hospitalized patients with severe or critical disease. In those patients, peak viral load in the upper respiratory tract occurs during the second week post onset of symptoms (POS), while viral clearance is achieved after 10 days in more than 90% of patients with mild disease [6]. Elevated cytokine levels, including IL-6 and IP-10, increased CRP and profound T-cell lymphopenia signal disease worsening [7-10]. This dysregulated inflammatory response is thought to result from an initial impairment of interferon production and thus reduced early viral control. Several reports have suggested that SARS-CoV-2 antibody titers are higher in patients with more severe disease [3, 11]. Activated SARS-CoV-2-specific CD4 and CD8 T cells have been reported in a few studies [12-14].

Yet viral shedding and immune responses in patients with mild COVID-19, the most frequent form of disease, remain poorly characterized. Comprehensive studies investigating viral shedding and immune responses at multiple time points in these patients are lacking. Available descriptions and interpretations of viral behavior and immunological responses in more severely ill patients may be biased by the rapid implementation of immunomodulatory treatments.

In our region, the first known case of COVID-19 was identified late February 2020. In line with public health policy early in the outbreak, every confirmed patient was hospitalized, including previously healthy young adults with only mild influenza-like illnesses. Here we report the detailed clinical, virological and immunological characterization of the first five patients assessed at our institution, from the day of diagnosis until convalescence.

## Materials and Methods

### Study population and assessment

The first five patients (P1-P5) with COVID-19 confirmed by real-time reverse transcriptase polymerase chain reaction (rtRT-PCR) and quarantined at HUG were included in this single-center prospective cohort. These patients were screened for SARS-CoV-2 according to the national guidelines in place at the time, which recommended screening travelers returning from Italy and reporting respiratory and/or flulike symptoms. Ethical approval was waived by the local ethics committee (BASEC Req-2020-00143); written informed consent for sample collection and coded data gathering was obtained from each patient. Clinical characteristics were described according to the Severe Emerging Infections clinical protocol of the International Severe Acute Respiratory and Emerging Infection Consortium [15], and disease severity was assessed according to WHO’s classification, where mild illness corresponds to mild symptoms without pneumonia [16]. If the patient agreed, we collected daily nasopharyngeal, oropharyngeal, conjunctival, sweat and anal swabs during hospitalization, as well as saliva, urine, and stool samples (see Supplementary methods). Plasma and serum were collected daily during hospitalization and at days 14 ±2 and 28 ±7 POS following discharge for viral load, antibody and cytokine quantification. Whole blood samples were used to isolate peripheral blood mononuclear cells (PBMC) to assess cellular responses.

### Virological assessment

Quantitative real-time RT-PCR was performed on all clinical samples. RNA was extracted using the NucliSens eMAG extraction (BioMérieux, France) and quantified with the Charité rtRT-PCR protocol [17] using *in vitro* transcribed RNA for quantification (European Virus Archive) [18]. Screening for viral co-infections was performed via a large in-house panel of rt(RT-)PCR (see supplement). Viral culture was performed on VeroE6 cells as previously described [19]. Positive isolation of SARS-CoV-2 was confirmed upon presence of cytopathic effect (CPE) and an increase in viral RNA between two consecutive supernatant samples. Nasopharyngeal swabs (NPS) from P1 to P4 were tested by high-throughput sequencing (HTS) analysis in the context of SARS-CoV-2 viral genome epidemiological surveillance in Switzerland. Each sample was treated as previously published (see supplement) [20].

### Measurement of inflammatory markers and blood cell phenotyping

Concentrations of 24 markers (see supplement) were measured in cryopreserved plasma using a Luminex assay (Magnetic Luminex assay, R&D Systems) according to the supplier’s instructions. For data points below the detection limit, a value of 50% of the last standard dilution value was assumed. For phenotyping of blood cells, cryopreserved PBMCs were thawed, counted and divided to perform T-cell and monocyte phenotyping. T cells were stained in PBS with LIVE/DEAD™ Stain Kit to exclude dead cells, anti-CD3, -CD4, -CD8, -CD38 and anti-HLA-DR antibodies and were then fixed and permeabilized. Anti-Granzyme B and anti-Ki67 antibodies were used for intracellular staining. For phenotyping of monocytes, cells were stained with FcR binding inhibitor, anti-CD3, -HLA-DR, -CD40, -CD123, -CD169, -CD20, -CCR2, -CD14, -CD16, -CD86, -CD163 and anti-CCR7 antibodies. All data were acquired the same day on a Fortessa II cytometer (BD Biosciences) and analyzed using FlowJo software (V10, Tree Star).

### Antibody assays

S1 domain-specific IgG and IgA antibody response was measured using a commercially available kit (Euroimmun AG, Lübeck, Germany, #EI 2606-9601 G and EI 2606-9601 A) according to the manufacturer’s instructions. To detect antibody against the complete S protein, ELISA plates were coated with a purified trimerized S protein kindly provided by the Ecole polytechnique fédérale de Lausanne (EPFL). IgG, IgA and IgM antibody titers were determined using a SARS-CoV-2 complete spike (S) protein-based rIFA [21, 22]. Neutralizing antibodies were quantified on VeroE6 cells infected with a VSV-based SARS-CoV-2 pseudotype expressing a 19-amino-acid, C-terminal truncated spike protein [23-25] (NCBI Reference sequence: NC_045512.2) with serially diluted sera. VeroE6 cells were infected with the virus-serum-mixture, and GFP positive infected cells were counted to assess titers [21]. Details can be found in the supplement.

## Results

### Clinical characteristics

Baseline demographic and clinical characteristics of the patients, all adult men, are shown in table 1. Four patients (P1-4) had only mild upper respiratory tract infections (URTI) and required no or only antipyretic treatment. Their presentation was typical for the majority of COVID-19 patients (sore throat, cough, fever), including P1, who had almost no symptoms. The fifth patient (P5) had a severe illness with bilateral pneumonia; computed tomography (CT) revealed bilateral patchy infiltrates involving each pulmonary lobe and a left posterior basal consolidation 7 days POS (Sup Figure 1). On day 3 of hospitalization, he developed a maculopapular rash on the back and the trunk. His treatment included oxygen support, a 7-day course of antibiotics and lopinavir/ritonavir [26]. He recovered and was discharged after 10 days (day 15 POS). All laboratory parameters and in-depth clinical descriptions are available in supplementary tables 1-6.

**Table 1:** Main characteristics of the patients

|  | Patient 1 | Patient 2 | Patient 3 | Patient 4 | Patient 5 |
| --- | --- | --- | --- | --- | --- |
| Age range | 20-29 | 30-39 | 50-59 | 20-29 | 60-69 |
| Sex | Male | Male | Male | Male | Male |
| Co-morbidities | None | None | None relevant for SARS-CoV-2 infection | None | None relevant for SARS-CoV-2 infection |
| Duration of Illness (days) | 1 | 5 | 6 | 9 | 16 |
| Maximal temperature (°C) * | 38 | 38.4 | 38.5 | 39 | 37.3 |
| Oxygene Saturation (RA/with oxygen (FiO2)) | 99% AA | 99% AA | 99% AA | 100% AA | 92% (32%) |
| Symptoms | Fever, rhinorrhea, headache | Fever, odynodysphagia, dry cough, headache | Fever, odynodysphagia, dry cough, chest pain | Fever, fatigue, dry cough | Fever**, fatigue, productive cough, skin rash |
| Diagnostic | Upper Respiratory Tract Infection | Upper Respiratory Tract Infection | Upper Respiratory Tract Infection | Upper Respiratory Tract Infection | Severe bilateral pneumonia |
| Treatment | None | Paracetamol | Alfuzosine, Paracetamol | Ibuprofen, Paracetamol, Enoxaparin | Co-amoxicilline + Clarythromycine, Piperacillin/Tazobactam, Lopinavir/ritonavir, Folic acid, Enoxaparin |

**Figure 1.**
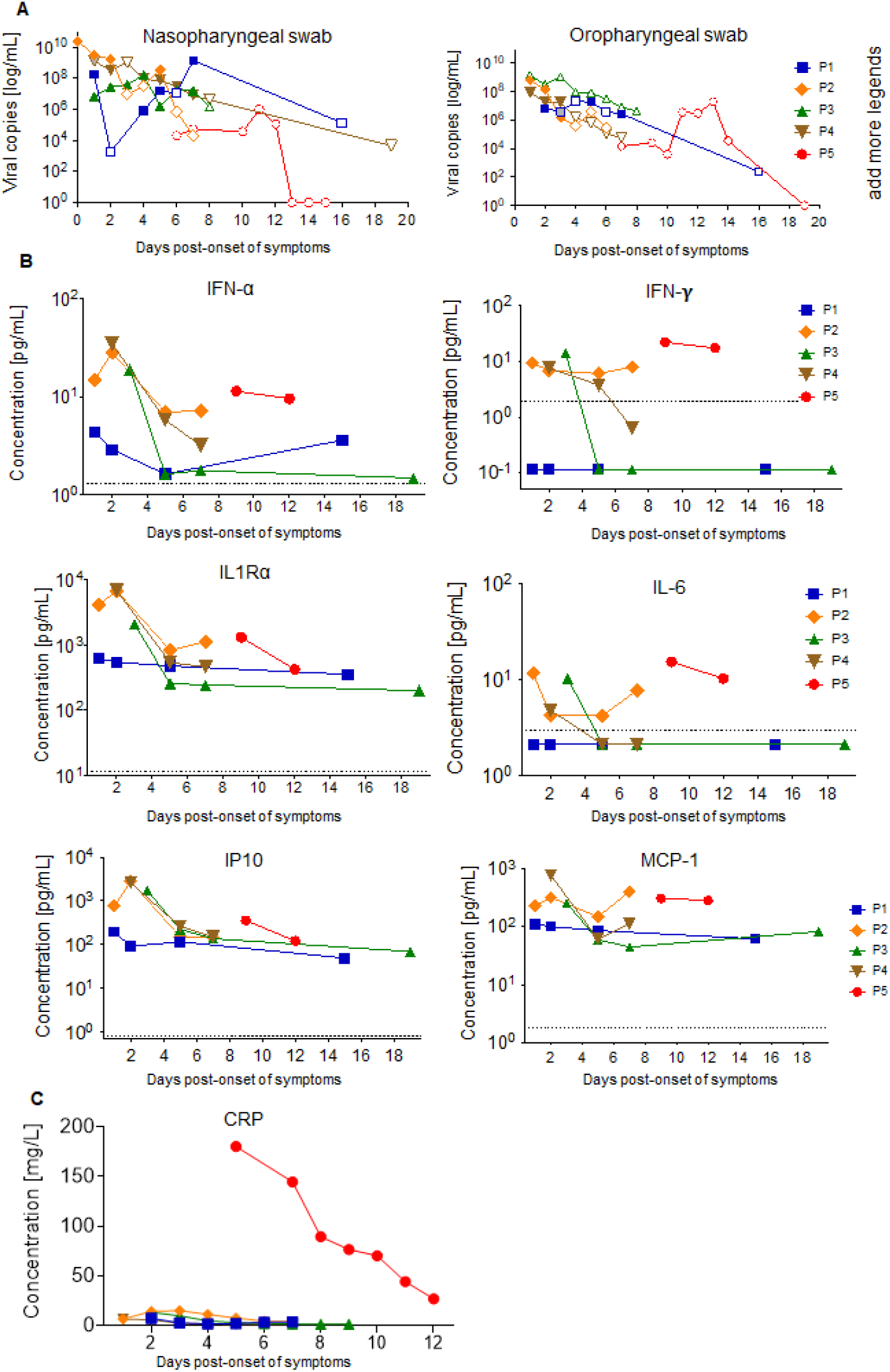
Kinetics of viral load and inflammatory markers. (A) Viral kinetics of nasopharyngeal (NPS) and oropharyngeal swab (OPS) samples. Plain logo: isolation of infectious virus in cell culture. Empty logo: infectious virus could not be isolated in cell culture. In grey: sample not available for virus isolation (B) Cytokine and chemokines dynamics. Individual concentrations in pg/mL for each marker were plotted at different days POS for each patient. The dotted line represents the limit of detection for each marker. Samples with undetectable concentrations were arbitrarily given a value of 50% of the last standard dilution value. (C) C-reactive protein (CRP) dynamics. Concentration in mg/L was plotted at different days POS for each patient.

### Viral shedding patterns

SARS-CoV-2 was detected by rtRT-PCR in both the oropharyngeal (OPS) and nasopharyngeal swabs (NPS) of each patient, with high viral loads (VL) that declined over time (Figure 1A). Higher VL were detected in NPS than OPS (mean of 1.1E9 viral RNA copies/mL and 4.1E7 copies/mL, respectively) with a peak viral load of 2.5E10 copies/mL for P2, sampled at the day of symptom onset. Lower viral loads in NPS, relative to viral loads of P1-4 on days 6-7 POS, were consistently observed for P5, for whom sampling started only on day 6 POS and covered the second week of illness.

Isolation of infectious virus was successful from both NPS and OPS during the first week of illness for the four mild cases, with the last positive virus isolation on day 7 for two patients (P1 and 3). The mean viral load in samples affording successful isolation was 1.2E9 copies/mL; the lowest RNA viral load in a sample from which a positive isolate could be obtained was 1.4E6 copies/mL. No infectious virus could be isolated from any sample from P5.

Prolonged RNA detection was observed in three patients, with persistent positive PCR results on days 14 (P5), 16 (P1) and 19 (P3) POS, albeit with low viral loads. RNA was also detected by PCR in saliva and stool specimens, with peak VLs of 7.8E6 and 8.6E5 copies/mL, respectively (Sup Figure S2). No virus was detected in other specimen types, and no RNAemia was observed in plasma. No virus isolation was attempted on non-respiratory samples.

Full genome sequences from P1-4 were found to cluster with sequences representative of the closest geographically related outbreak at that time, which was in Northern Italy. Molecular epidemiology is consistent with histories of travel to the region shortly before symptom onset for P1, P2 and P3 (Figure S3). A co-infection with adenovirus at a threshold cycle (Ct) value of 36 was detected in P1 on NPS collected at the time of his diagnosis. No other co-infections were detected in P2-P5.

### Kinetics of the innate response

An increase in cytokine levels was observed in the first days POS (Figure 1B). Patients with mild symptoms had an increase in IFN-α, IFN-γ and IP-10 around day 2-3, which returned to baseline as symptoms resolved and VL decreased. IL-6, which has been associated with severe disease [27], was slightly more elevated at early time points, as were IL-1Ra and MCP-1. Their levels were similar to the ones observed in P5 when he was oxygen-dependent. Cytokine levels were generally lower in the patient with the mildest symptoms (P1). There were no significant changes in the other cytokines tested, such as TNFα, IL-1β, IL-2 or IL-8 (not shown). CRP levels were much higher in P5 than in patients with mild disease (Figure 1C), suggesting that innate responses can be efficiently triggered in mild disease.

In line with the cytokine data, total numbers of monocytes and neutrophils were higher at early time points relative to later time points (Tables S2-S6). Because monocytes and macrophages have been shown to play an important role in pathogenesis [28], we performed a deeper phenotypic analysis of the circulating monocyte population by flow cytometry; it revealed a population of intermediate monocytes (CD14^+^ CD16^high^ cells), accounting for the majority of the monocytes detected in peripheral blood at days 2-3 POS (3% to 36% of cells from the live gate) (Figure 2A, B). This was seen in all patients irrespective of symptom severity, and decreased for one week POS in all mild patients with sequential samples. Their number was higher in P5 at the two time points tested. Intermediate monocytes expressed the highest levels of activation markers (CD86 and CD40) at 2-3 days POS, which was followed by the peak in the expression of differentiation and migration markers (HLA-DR, CD169, CD163, CCR-7) at one week POS (Figure 2C). In summary, we observed activation of innate responses in all patients, with a clear increase in type I interferon and pro-inflammatory cytokines, as well as significant increase in intermediate monocytes with activation, differentiation and migration patterns.

**Figure 2.**
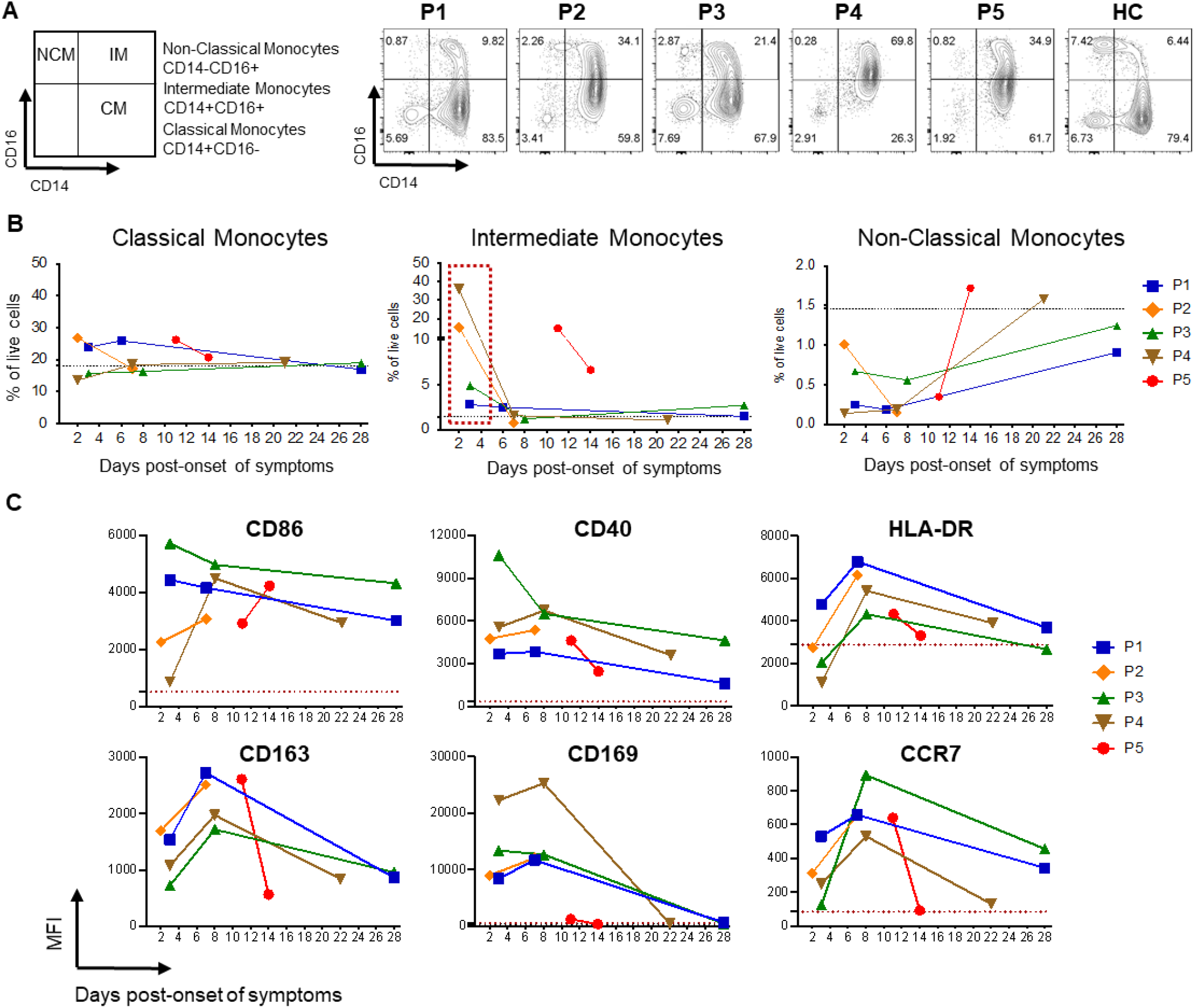
Early increase in intermediate monocytes after Sars-CoV-2 infection. (A) Gating strategy for the identification different monocytes subsets: classical monocytes (CM), intermediate monocytes (IM), non-classical monocytes (NCM) by flow cytometry after gating out dead cells and lineage (CD3, CD20) cells, and gating on HLA-DR+ cells. Early time points flow cytometry counterplots from P1 to P5 as well as a healthy control (HC) are shown. (B) Percentage of CM, IM and NCM calculated from the lymphocyte live gate for each patient at different days POS. The dotted line represents one healthy donor. (C) Geometric mean (GM) MFI value for different activation and migration markers in Intermediate Monocytes. The dotted line in red shows the mean of the isotype controls for each marker.

### Antibody and cellular responses

Serum Ab titers against the spike protein (complete spike and S1 domain) were evaluated by ELISA and rIFA assay (Figure 3). Patients 1, 3, 4 and 5 seroconverted by two weeks POS, including P1 who had very mild and transient symptoms. For P2, no sample was available beyond day 7. P3 had no detectable IgA or IgM, and only low IgG levels (Figure 3A and B). Interestingly, this patient seroconverted only 3 weeks POS in S1-based ELISA (Figure 3A), but had already seroconverted after two weeks in complete S-based assays (rIFA and ELISA) (Figure 3A and B) and in neutralization assays (Figure 3C), indicating a potential role of S2-domain-specific antibodies in virus neutralization. P5 had the highest antibody levels and seroconverted earlier than patients with mild symptoms. Antibodies from all patients except P2, for whom samples from later time points were not available, were able to neutralize SARS-CoV-2 S-pseudotyped viruses; P5 again demonstrated the highest neutralizing antibody titers (Figure 3C). For P1 and P3 a late serum sample was obtained at 69 days POS. He had already showed a decline in IgG, IgA, IgM and neutralizing Ab titers (Figure 3A-C) while antibody levels of P3 declined only minimally (rIFA, S1 ELISA) or stayed the same (NT).

**Figure 3.**
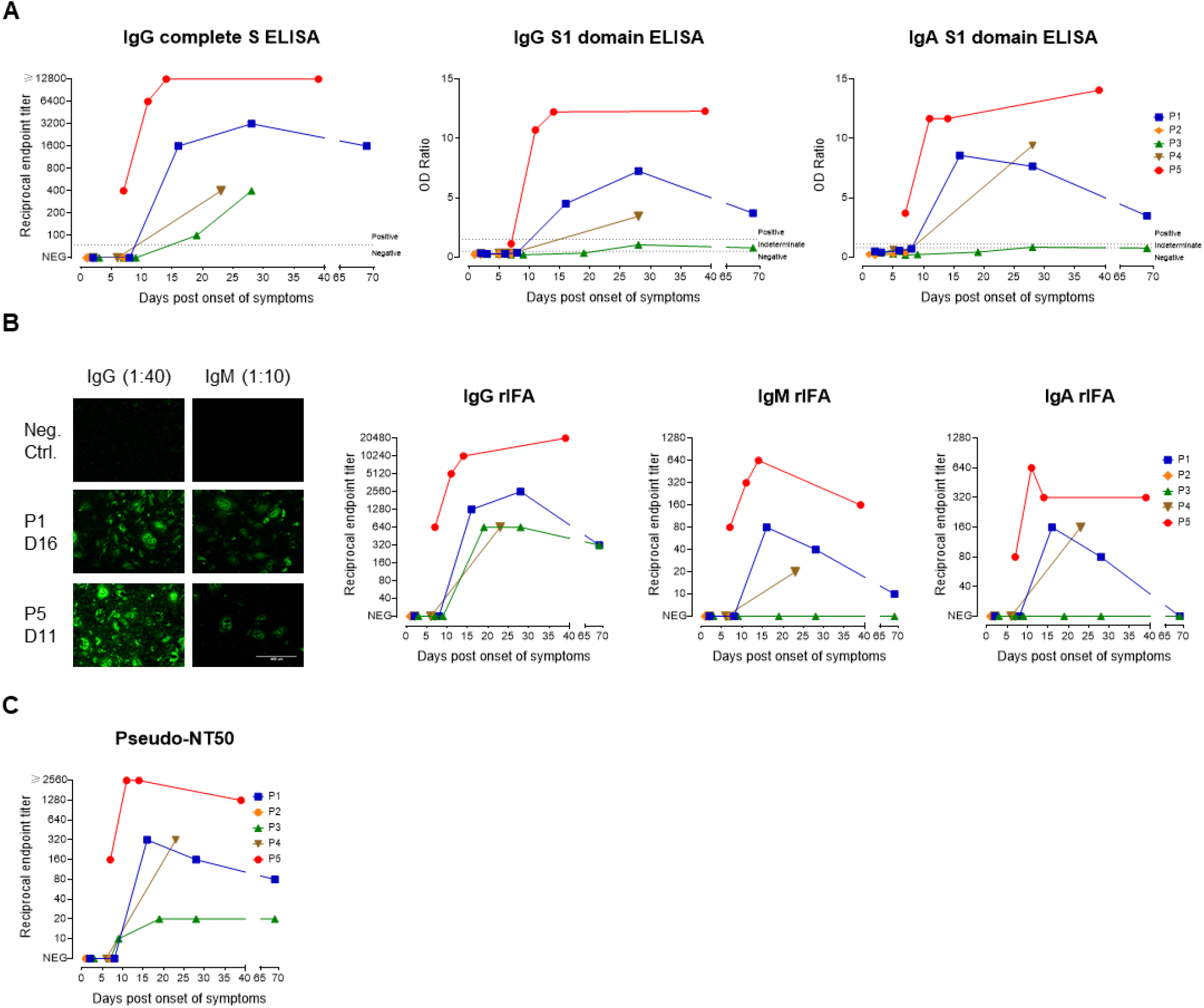
Antibody response to SARS-CoV-2. (A) IgG and IgA antibody kinetics for P1-5 using ELISA assays with either SARS-CoV-2 full S (only IgG, left graph) or S1-domain (IgG and IgA, middle and right graph) as antigen. For Full S ELISA endpoint titers were determined (starting dilution 1:100) whereas for S1-based ELISA the ratio of sample/calibrator was measured (dilution 1:100). (B) Antibody kinetics using a recombinant full S-based immunofluorescence assay (rIFA) for IgG, IgM and IgA isotypes using starting dilution of 1:40 (IgG and IgA) or 1:10 (IgM). The left-panel shows representative rIFA staining patterns for IgG and IgM for two patients and a negative control serum. (C) Kinetics of neutralizing antibody endpoint titers using a pseudotyped VSV neutralization assay (starting dilution 1:10).

We measured the frequency of T lymphocytes and detected a marked reduction in total lymphocytes, particularly of CD8+ T cells, in P5 (Figure 4A, B). We further characterized the T-cell phenotype in terms of activation (expression of markers CD38 and HLA-DR), proliferation (Ki-67) of CD4 and CD8 T cells, and the expression of Granzyme B for CD8 T cells (Figure 4C, D). The highest frequency of activated CD8 T cells was detected in P5, with very few proliferating cells (Figure 4C). Although the frequency of activated CD8 T cells was lower in patients with mild symptoms, activated CD8 T cells expressed Granzyme B in all patients. For CD4 T cells, the frequency of activated cells was higher in P5 (CD38+HLA-DR+ CD4 T cells 2.8%) than in healthy individuals (mean 0.49%), and even in patients with mild symptoms (CD38+HLA-DR+ CD4 T cells <1.35%). Activated CD4 T cells were proliferating in all infected patients (Figure 4D). Altogether, the detection of activated and proliferative T cells in all patients suggests that even patients with mild disease are able to mount a T-cell-mediated immune response to SARS-CoV-2.

**Figure 4.**
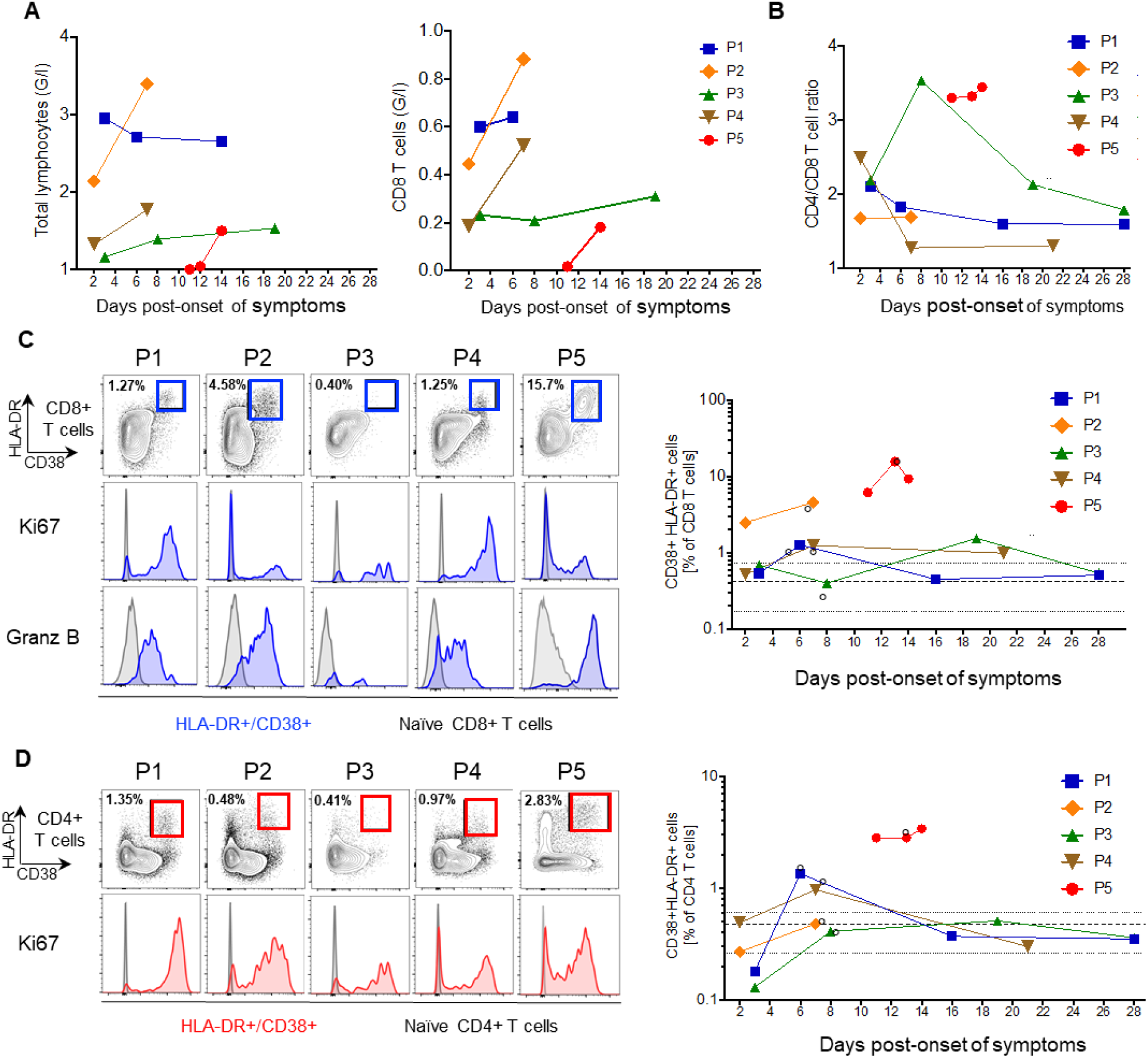
Kinetics of T-cell immune responses. (A) Graphs show the frequency of total lymphocytes and of CD8 T cells. (B) The ratio of CD4/CD8 T cells was determined by flow cytometry as the ratio of the frequencies of CD4 and CD8 T cells (percentage of CD3 T cells) (C) Representative flow plots are shown for CD8 T cell activation markers CD38 and HLA-DR. Histograms are shown for the expression of Ki67 (middle) and Granzyme B (lower) of activated (blue) and naive CD8 T cells (gray, gated on CD45RA+ CCR7+ CD8 T cells). The summary graph depicts the kinetics of HLA-DR + CD38+ CD8 T cells at different time points after symptoms onset. The bold dotted line is the mean of three healthy controls with its range (thin dotted line); “°” identifies the time points that are shown as flow plots. (D) Similar to C, representation of flow plots HLA-DR+CD38+ CD4 T cells (red), expressing the proliferation marker Ki67, with summarized data in the right graph.

## Discussion

We report the kinetics of viral load and immune responses in the first five COVID-19 patients hospitalized in our institution presenting with mild (P1-4) and severe (P5) disease. The most striking finding was a robust innate response in patients with mild symptoms characterized by early activation of monocytes. We also found prolonged viral RNA detection and measurable SARS-CoV-2 specific cellular and Ab responses despite the paucity of symptoms.

We documented high VL in the upper respiratory tract at diagnosis and the presence of infectious virus in both NPS and OPS, in samples taken hours POS in patients presenting with mild illness. Similar results have been documented in the pre-symptomatic phase [29] and can partly explain the high transmissibility of COVID-19. In all patients with mild disease, infectious virus could be isolated several days after symptom onset, up to day 7. Notably, symptoms had already waned for 5 days before the last successful isolation of infectious virus P1. We could not isolate SARS-CoV-2 in clinical specimens containing less than 1.4E6 viral RNA copies/ml, in line with other reports [22]. Interestingly, no virus could be isolated from the NPS during the second week of illness, in the patient with the severe disease, possibly indicating antibody-mediated viral neutralization at that time. Except in stools and saliva, viral RNA was observed inconsistently and at low levels in other body fluids. This suggests that these fluids are not a major source of transmission. While patients with mild symptoms tend to have more limited shedding [6], prolonged RNA detection has also been reported by others [30]. In our study, no infectious virus could be retrieved from such samples, arguing for lingering remnant RNA rather than for active replication. While there is a need to confirm these findings in larger cohorts, they provide important information for discharge criteria and public-health decisions regarding isolation. Our data on isolation of infectious virus support most recommendations currently in effect, which advise isolation for at least 10 days POS for infection control, even in patients with very brief symptom durations.

Monocytes and macrophages play a critical role in the immunopathology of COVID-19 [28]. An increased number of cytokine-producing CD14+CD16+ monocytes and activated macrophages are seen in the blood and broncho-alveolar lavages of severe patients, respectively [31-33]. Zhou et al showed that increases in activated intermediated monocytes were mainly seen in intensive-care-unit (ICU) patients as opposed to non-ICU patients [34]. An interesting observation is that the rapid and drastic impact of SARS-CoV-2 infection on circulating monocytes in patients with very mild symptoms has a profile that is similar to what is reported by Zhou and colleagues in ICU patients. The new observation could be due to the fact that our study captured very early responses. The rapid mobilization of monocytes is indicative of a robust local antiviral response. In patients without complications, this early mobilization of monocytes is not sustained and rapidly declines to baseline one-week POS. This monocyte signature is not unique to SARS-CoV-2: it has been described for dengue fever [35, 36] and other viral diseases [10, 37, 38].

In our study, all patients with mild symptoms, and with samples available from day 7 POS or later, mounted adaptive immune responses leading to neutralizing Ab. The question of the durability of the response and its quality over time remains open. As observed for P1 but not for P3, Ab titers including neutralizing Abs may rapidly decline in some non- or pauci-symptomatic COVID-19 patients. Experimental studies showed that immunity against reinfection by other human CoVs wanes over a few months, enabling reinfection within the first year after acute disease [39]. Low levels of non-neutralizing Ab may enhance disease severity at time of re-exposure [40], through an Ab-mediated mechanism described for other viruses. This could have important implications for both the clinical severity of repeat exposure and vaccination strategies eliciting more binding than neutralizing antibodies [41].

Our study has limitations. Although we were able to perform a comprehensive analysis of the immune response and viral shedding, the description is limited to five patients (with early time points missing for P5, and later time points missing for P2) and our interpretation of the association between viral load, innate and adaptive responses in patients with mild COVID-19 requires confirmation with a larger sample size. Finally, we were unable to follow all patients for late time points and cannot reach a conclusion about the durability of SARS-CoV-2 specific immunity according to disease pattern.

## Conclusion

We provide a comprehensive immunological and virological profile of the first five patients diagnosed in our institution. Four had only mild and/or very transient symptoms and none received immunomodulatory treatments. Importantly we found that in both patients with mild symptoms and prolonged high viral loads, as well as one with severe disease, no virus could be isolated after the first week of illness.

Even in patients with mild symptoms, we found a robust innate response characterized by the mobilization of activated monocytes in the first days of infection, associated with high viral loads and the induction of interferon-dependent circulating cytokine responses. All patients mounted SARS-CoV-2 specific adaptive responses including neutralizing antibodies.

## Data Availability

All data referred to in the manuscript are stored at Geneva University Hospitals.

## Table legend

Table 1: Main characteristics of the patients

RA: room air

*: during hospitalization

**: self-measured, at home, before hospitalization

## Author Contributions

PV, IE, LK, and CE designed the study. PV, AC, DLV, MS and FJ collected clinical data and samples. FP, GT and IE conducted the virological analysis. SC and FL performed HTS analysis. CE, BM, PAM, SL, AMD and CAS conducted immunological analysis. PV, BM, CE, SC, AMD and IE contributed to the writing of the manuscript. All authors critically reviewed the manuscript and accepted the final version.

## Acknowledgements

We thank Erik Boehm for editorial assistance, Sabine Yerly and Manel Essaidi-Laziosi for technical assistance, Angela Huttner for her meaningful insights and to colleagues at the protein core facility at the Ecole Polytechnique Fédérale de Lausanne for providing the complete S protein. We are also grateful to the patients who have agreed to participate in this study.

## Supplementary appendix

### Supplementary Methods

#### Sample collection

We collected daily nasopharyngeal, oropharyngeal, conjunctival, sweat and anal swabs as well as saliva, urine and stool samples if the patient agreed, using 3mL universal transport medium tubes (Copan, Brescia, Italy). Urine and stools were collected daily (when available) in plastic tubes without additives. Plasma and serum were collected in EDTA and in SST II plastic tubes, daily during hospitalization and at days 14 +/-2 and 28 +/-7 post onset of symptoms (POS) following discharge for viral load, antibody (Ab) and cytokine quantification. Cell-preparation tubes (CPT) with sodium citrate were used for collection of whole blood and separation of peripheral blood mononuclear cells (PBMC) to assess cellular responses.

#### Assessment of infectious viruses

VeroE6 cells were seeded at a density of 8×10^4^ cells/well in a 24-well plate and inoculated with 200 μl of viral transport medium the following day. Cells were inoculated for 1h at 37°C, then inoculum was removed, cells were washed 1x with PBS and then regular cell growth medium containing 10% FCS was added. Cells were observed on days 2, 4, and 6 for the presence of CPE by light microscopy. Supernatant was harvested upon the first observation of a CPE, or, if no CPE was observed, at the end of the experiment on day 6.

#### Testing for viral co-infections

Nasopharyngeal swabs were screened by acid nucleic detection for the presence of influenza A and B virus, respiratory syncytial virus (RSV) A and B, parainfluenza 1 to 4, human metapneumovirus, rhinoviruses, enteroviruses, bocavirus 1, adenovirus and human coronaviruses 229E, OC43, HKU1 and NL63 as previously described. (1)

#### High-throughput sequencing

110 µl of NPS were centrifuged at 10,000 × g for 10 min. One-hundred μl of cell-free supernatant were treated with 20 U of Turbo DNAse (Ambion, Rotkreuz, Switzerland). Viral nucleic acids were extracted with TRIzol (Invitrogen,Carlsbad, CA, USA) and resuspended in 10 μl of RNAse-free water. Ribosomal RNA was removed using the Ribo-Zero Gold depletion kit (Illumina, San Diego, US). Thereafter, libraries were generated using the TruSeq total RNA preparation protocol (Illumina) with dual indexing and loaded on the HiSeq 4000 platform (Illumina) using the 2×100-bp protocol. Raw data were analysed as follows: duplicate reads were removed using cd-hit (v4.6.8). Then reads were then trimmed to remove low-quality and adapter sequences using Trimmomatic (v0.33). Next reads were mapped against the reference sequences MN908947.3 using the SNAP nucleotide aligner program. (2) The four hCoV-19 complete sequences hCoV-19/Switzerland/GE3895/2020 (P1), hCoV-19/Switzerland/GE9586/2020 (P2), hCoV-19/Switzerland/GE3121/2020 (P3), and hCoV-19/Switzerland/GE0199/2020 (P4) were all submitted and made available via GISAID.

#### Assessment of innate imunity

The following list of markers were tested in Luminex: CD40L, GM-CSF, Granzyme B, IFN-a, IFN-g, IL1a, IL1-b, IL1R-a, IL-2, IL-4, IL-6, IL-8, IL-10, IL-12p70, IL-13, IL-15, IL17A, IL-33, IP-10, MCP-1, MIP-1a, MIP-1b, PD-L1, TNF-a. The mean fluorescence intensity of each marker was read on the Bio-Plex 200 array reader (Bio-Rad Laboratories) using the Luminex xMAP Technology (Luminex Corporation). Sample concentrations were calculated using a five-parameter logistic regression curve (Bio-Plex Manager 6.0)

#### References for reagents used for cell phenotyping

LIVE/DEAD™ Fixable Aqua Dead Cell Stain Kit (Life Technologies), Fixation/Permeabilization kit (Invitrogen 00-8333-56), anti-CD3 (clone SK7 BioLegend), anti-CD4 (clone RPA-T4, BD), anti-CD8 (clone SK1, BD), CD38 (clone HIT2, BioLegend), HLA-DR (clone L243, BioLegend), Granzyme B (clone GB11, BD), Ki67 (clone B56, BD), FcR binding inhibitor (14-9161-73, Invitrogen), anti-CD3 (clone SP34-2, BD), HLA-DR (clone G46-6, BD), CD40 (clone 5C3, BD), CD123 (clone 7G3, BD), CD169 (clone 7-239, Biolegend), CD20 (clone, BD), CCR2 (clone REA264, Miltenyi), CD14 (clone M5E2, BD), CD16 (clone 3G8, BD), CD86 (clone IT2.2 Biolegend), CD163 (clone GHI/61, BD), CCR7 (clone 3D12, BD)

#### Complete S protein-based ELISA

We coated 37.5ng/well of a purified trimerized S protein diluted in 0.1M sodium carbonate buffer pH 9.6 O/N at 4°C. Plates were blocked (5% milk PBS-T) and 100 ul of 2-fold serially diluted sera (range 100-12,800) were applied to wells with and without antigen. After incubation (1h at 37°C) and washing (3x 1min with PBS-T) 100ul HRP-conjugated anti-human IgG (Jackson Immunoresearch, #109-036-098) diluted 1:16,000 was added. After incubation plates were washed and 100ul of TMB substrate (Invitrogen) was added for 10min. Reaction was stopped by 100ul of 1N HCl and plates were read at 450nm. OD450 values of blank wells were subtracted from values of antigen containing wells for each serum and each dilution. The cut-off to determine the ELISA titer was set at 0.29 OD450, just before OD values reach the plateau phase.

#### rIFA assay

Briefly, pCG1 vector expressing SARS-CoV-2 S protein (kindly provided by M. Hoffmann and S. Pöhlmann, DPZ Göttingen) was transfected into Vero B4 cells using Fugene HD (Promega #E2311), spotted on multitest microscopy slides (DUNN Labortechnik GmbH #40-412-05) and fixed with ice-cold Acetone/Methanol (1:1). To perform the rIFA sera were inactivated for 30min at 56°C. To remove IgG antibodies for determination of IgM antibody titres, sera were treated with Eurosorb reagent (Euroimmun AG #1270-0145). Slides were rehydrated in PBS-T for 5min and blocked (5% milk PBS-T) for 30min at RT. Sera were diluted using a starting dilution of 1:40 for IgG/IgA and 1:10 for IgM and 30μl were applied to each spot. After incubation (1h, 37°C) slides were washed (3×1min, PBS-T) and 25μl of Alexa488-conjugated goat anti-human-IgG, -IgM. or -IgA antibody (Jackson ImmunoResearch #109-036-098, #, #) diluted 1:200 in PBS was applied. After incubation (37°C, 45min), slides were washed as before and briefly rinsed with dH_2_O before mounting with glycerol.

#### Quantification of neutralizing antibodies

VeroE6 cells were seeded in 96-well plates at 2 x 10^4^ cells per well and grown into confluent monolayer overnight. Sera from patients were inactivated at 56°C for 30 minutes and diluted from 1:5 to 1:1280 in DMEM 2% FBS. VSV-based SARS-CoV-2 pseudotypes (3, 4) expressing a 19 amino acids C-terminal truncated spike protein (5) (NCBI Reference sequence: NC_045512.2) were diluted in DMEM 2% FCS in order to have MOI=0.01 per well and added on top of serum dilutions (final serum dilutions obtained were from 1:10 to 1:2560). The virus-serum mix was incubated at 37°C, for 2h. Vero E6 were then infected with 100µl of virus-serum mixtures. After incubation at 37°C for 1.5h, cells were washed once with 1X PBS and DMEM 10% FBS was added. After 16-20h of incubation at 37°C, 5% CO_2_ cells were fixed with 4% formaldehyde solution for 15min at 37°C and nuclei stained with 1µg/ml DAPI solution. GFP positive infected cells were counted with ImageXpress® Micro Widefield High Content Screening System (Molecular Devices) and data analyzed with MetaXpress 5.1.0.41 software.

### Supplementary material

**Supplementary Table 1:** Detailed characteristics of the patients Legend: CRP: C Reactive Protein; PCT: procalcitonin; AST = aspartate aminotransferase; ALT = alanine aminotransferase; GGT = gamma-glutamyl transferase

|  | Patient 1 | Patient 2 | Patient 3 | Patient 4 | Patient 5 |
| --- | --- | --- | --- | --- | --- |
| **Smoking history** | Occasional smoker | Current smoker | No | No | No |
| **Epidemiological exposure** | Travelled to country with local transmission | Travelled to country with local transmission | Travelled to country with local transmission | Relative of patient 1 | Unclear; met asymptomatic relatives back from countries with local transmission 15 and 12 days before symptoms onset |
| **Other** | *Chlamydophila/Myco plasma* (Throat swab) PCR neg | Blood culture on admission: no pathogen growth | Urine culture: no pathogen growth | Blood culture on admission: no pathogen growth | Blood culture on admission: no pathogen growth<br>Urinary Legionella pneumophila and Streptococcus pneumoniae Ag negative<br>Chlamydophila/Myco plasma (Throat swab) PCR neg<br>Urine culture: no pathogen growth<br>Skin biopsy : suprabasal acantholysis, ballooned multinuclear cells, lymphohistiocytic infiltrate of the dermis |
| **Viral co-infection** | Adenovirus | None detected | None detected | None detected | None detected |
| **Imaging** |  |  |  |  |  |
| **Chest X-ray** | N/A | N/A | N/A | No infiltrates | Bilateral patchy infiltrates |
| **CT-Scan** | N/A | N/A | N/A | N/A | Left postero-basal consolidation, bilateral patchy infiltrates |

**Table S2:**
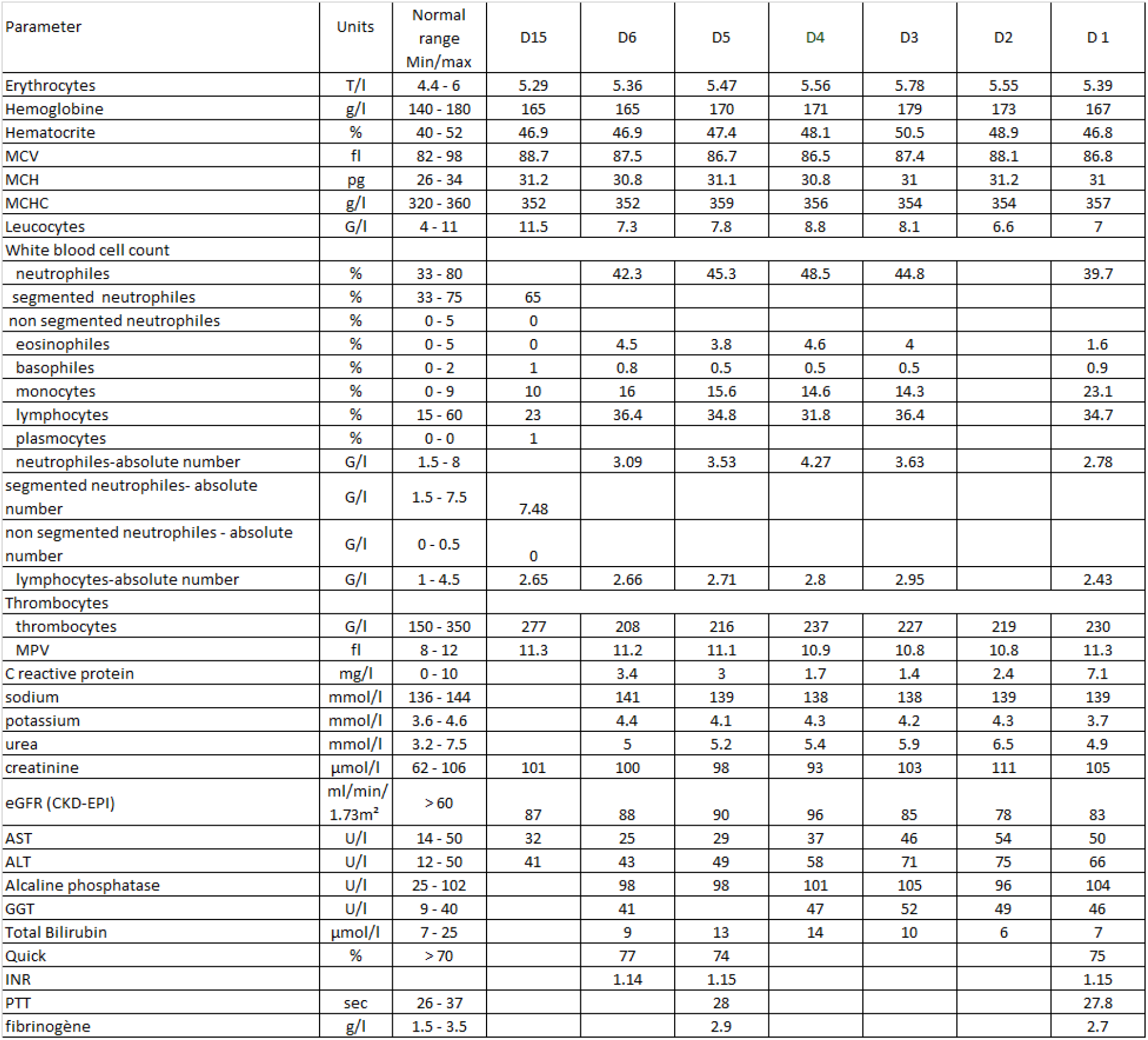
Patient 1’s laboratory parameters during hospitalization

**Table S3:** Patient 2’s laboratory parameters during hospitalization

| Parameter | Units | Normal range<br>Min/max | D7 | D6 | D5 | D4 | D3 | D2 | D1 |
| --- | --- | --- | --- | --- | --- | --- | --- | --- | --- |
| Erythrocytes | T/l | 4.4 - 6 | 5.48 | 5.45 | 5.4 | 5.52 | 5.38 | 5.51 | 5.64 |
| Hemoglobine | g/l | 140 - 180 | 158 | 158 | 157 | 160 | 157 | 159 | 167 |
| Hematocrite | % | 40 - 52 | 45.5 | 46 | 45.4 | 46.2 | 44.8 | 45.7 | 47.2 |
| MCV | fl | 82 - 98 | 83 | 84.4 | 84.1 | 83.7 | 83.3 | 82.9 | 83.7 |
| MCH | pg | 26 - 34 | 28.8 | 29 | 29.1 | 29 | 29.2 | 28.9 | 29.6 |
| MCHC | g/l | 320 - 360 | 347 | 343 | 346 | 346 | 350 | 348 | 354 |
| Leucocytes | G/l | 4 - 11 | 7.8 | 6.6 | 5.9 | 7.4 | 10.2 | 11.5 | 10.9 |
| White blood cell count |  |  |  |  |  |  |  |  |  |
| neutrophiles | % | 33 - 80 | 44.5 | 41.4 | 32.9 | 44.9 | 60.9 | 67.9 | 69 |
| segmented neutrophiles | % | 33 - 75 | 2.4 | 1.8 | 2.2 | 1.1 | 0.3 | 2.7 | 0.1 |
| non segmented neutrophiles | % | 0 - 5 | 0.5 | 0.5 | 0.5 | 0.3 | 0.2 | 0.3 | 0.4 |
| eosinophiles | % | 0 - 5 | 9.1 | 10.3 | 11.1 | 11.4 | 10.2 | 10.5 | 12.2 |
| basophiles | % | 0 - 2 | 43.5 | 46 | 53.3 | 42.3 | 28.4 | 18.6 | 18.3 |
| monocytes | % | 0 - 9 | 3.47 | 2.73 | 1.94 | 3.32 | 6.21 | 7.81 | 7.52 |
| lymphocytes | % | 15 - 60 | 3.39 | 3.04 | 3.14 | 3.13 | 2.9 | 2.14 | 1.99 |
| neutrophiles-absolute number | G/l | 1.5 - 8 | 214 | 204 | 189 | 176 | 131 | 194 | 244 |
| segmented neutrophiles- absolute number | G/l | 1.5 - 7.5 | 10.4 | 10.3 | 10.5 | 10.4 | 11.1 | 10.1 | 10.2 |
| non segmented neutrophiles - absolute number | G/l | 0 - 0.5 | 0 |  |  |  |  |  |  |
| lymphocytes-absolute number | G/l | 1 - 4.5 | 2.65 | 2.66 | 2.71 | 2.8 | 2.95 |  | 2.43 |
| Thrombocytes |  |  |  |  |  |  |  |  |  |
| thrombocytes | G/l | 150 - 350 | 277 | 208 | 216 | 237 | 227 | 219 | 230 |
| MPV | fl | 8 - 12 | 11.3 | 11.2 | 11.1 | 10.9 | 10.8 | 10.8 | 11.3 |
| C reactive protein | mg/l | 0 - 10 | 3.8 | 4.3 | 7.3 | 11 | 14.7 | 14 | 6.3 |
| sodium | mmol/l | 136 - 144 | 139 | 140 | 139 | 140 | 136 | 135 | 137 |
| potassium | mmol/l | 3.6 - 4.6 | 4.1 | 4.4 | 4.2 | 4.1 | 3.8 | 3.6 | 3.9 |
| urea | mmol/l | 3.2 - 7.5 | 3.6 | 4.2 | 3.7 | 4.2 | 4.2 | 3.9 | 5.2 |
| creatinine | μmol/l | 62 - 106 | 83 | 93 | 99 | 89 | 94 | 108 | 105 |
| eGFR (CKD-EPI) | ml/min/1.73m<sup>2</sup> | > 60 | 109 | 95 | 88 | 100 | 93 | 79 | 82 |
| AST | U/l | 14 - 50 | 26 | 29 | 29 | 29 | 30 | 37 | 39 |
| ALT | U/l | 12 - 50 | 44 | 45 | 41 | 41 | 47 | 52 | 58 |
| Alcaline phosphatase | U/l | 25 - 102 | 38 | 40 | 38 | 38 | 38 | 43 | 44 |
| GGT | U/l | 9 - 40 | 91 | 92 | 90 | 92 | 97 | 102 | 105 |
| Total Bilirubin | μmol/l | 7 - 25 | 8 | 7 | 6 | 7 | 7 | 7 | 7 |
| Quick | % | > 70 | >100 |  | >100 |  |  |  | 94 |
| INR |  |  | 1 |  | 1 |  |  |  | 1.03 |
| PTT | sec | 26 - 37 | 25.4 |  |  |  |  |  |  |

**Table S4:** Patient 3’s laboratory parameters during hospitalization

| Parameter | Units | Normal<br>range<br>Min/max | D19 | D9 | D8 | D7 | D6 | D5 | D4 | D3 | D2 |
| --- | --- | --- | --- | --- | --- | --- | --- | --- | --- | --- | --- |
| Erythrocytes | T/l | 4.4 - 6 | 4.7 | 4.84 | 4.66 | 4.96 | 5.06 | 4.96 | 4.83 | 5.1 | 4.69 |
| Hemoglobine | g/l | 140 - 180 | 138 | 141 | 140 | 142 | 147 | 146 | 143 | 151 | 140 |
| Hematocrite | % | 40 - 52 | 38.2 | 39.6 | 38.2 | 40.1 | 41.3 | 41 | 40.3 | 42.1 | 38.9 |
| MCV | fl | 82 - 98 | 81.3 | 81.8 | 82 | 80.8 | 81.6 | 82.7 | 83.4 | 82.5 | 82.9 |
| MCH | pg | 26 - 34 | 29.4 | 29.1 | 30 | 28.6 | 29.1 | 29.4 | 29.6 | 29.6 | 29.9 |
| MCHC | g/l | 320 - 360 | 361 | 356 | 366 | 354 | 356 | 356 | 355 | 359 | 360 |
| Leucocytes | G/l | 4 - 11 | 4.6 | 2.9 | 3.5 | 2.8 | 3 | 2.5 | 2.2 | 2.7 | 2.8 |
| White blood cell count |  |  |  |  |  |  |  |  |  |  |  |
| neutrophiles | % | 33 - 80 | 54.5 | 48 | 47.8 | 47.3 | 42.3 | 43 |  |  | 48.8 |
| segmented neutrophiles | % | 33 - 75 |  |  |  |  |  |  | 47 | 40 |  |
| non segmented neutrophiles | % | 0 - 5 |  |  |  |  |  |  | 0 | 8 |  |
| eosinophiles | % | 0 - 5 | 1.5 | 2.4 | 2.3 | 2.5 | 2.6 | 2 | 2 | 1 | 0.7 |
| basophiles | % | 0 - 2 | 0.7 | 0.3 | 0.3 | 0.4 | 0.3 | 0.4 | 0 | 0 | 0.7 |
| monocytes | % | 0 - 9 | 10 | 11.3 | 9.9 | 10.2 | 10.6 | 10 | 3 | 8 | 17.8 |
| lymphocytes | % | 15 - 60 | 33.3 | 38 | 39.7 | 39.6 | 44.2 | 44.6 | 48 | 43 | 32 |
| neutrophiles-absolute number | G/l | 1.5 - 8 | 2.51 | 1.39 | 1.67 | 1.32 | 1.27 | 1.08 |  |  | 1.37 |
| segmented neutrophiles- absolute number | G/l | 1.5 - 7.5 |  |  |  |  |  |  | 1.03 | 1.08 |  |
| non segmented neutrophiles - absolute number | G/l | 0 - 0.5 |  |  |  |  |  |  | 0 | 0.22 |  |
| lymphocytes-absolute number | G/l | 1 - 4.5 | 1.53 | 1.1 | 1.39 | 1.11 | 1.33 | 1.12 | 1.06 | 1.16 | 0.9 |
| Thrombocytes |  |  |  |  |  |  |  |  |  |  |  |
| thrombocytes | G/l | 150 - 350 | 184 | 132 | 117 | 122 | 128 | 130 | 114 | 132 | 116 |
| MPV | fl | 8 - 12 | 10.6 | 11.1 | 10.7 | 10.4 | 10.3 | 10.8 | 10.8 | 10.5 | 11 |
| C reactive protein | mg/l | 0 - 10 |  | 1 | 1.1 | 1.3 | 1.9 | 2.7 | 4.5 | 9.6 | 13.7 |
| sodium | mmol/l | 136 - 144 |  | 139 | 139 | 140 | 142 | 141 | 141 |  | 143 |
| potassium | mmol/l | 3.6 - 4.6 |  | 4.2 | 4.4 | 4.3 | 4.3 | 4 | 4.3 |  | 4 |
| urea | mmol/l | 3.2 - 7.5 | 5.2 | 4.8 | 5.6 | 4.9 | 4.7 | 4.8 | 4.8 | 5.6 | 5.6 |
| creatinine | µmol/l | 62 - 106 | 98 | 82 | 87 | 72 | 76 | 76 | 75 | 85 | 82 |
| eGFR (CKD-EPI) | ml/min/1.73m² | > 60 | 75 | 92 | 86 | 100 | 98 | 98 | 98 | 89 | 92 |
| AST | U/l | 14 - 50 | 30 | 39 | 36 | 45 | 60 | 29 | 23 | 32 | 26 |
| ALT | U/l | 12 - 50 | 34 | 77 | 76 | 87 | 96 | 32 | 24 | 30 | 27 |
| Alcaline phosphatase | U/l | 25 - 102 |  | 59 | 50 | 47 | 50 | 51 | 56 | 60 | 56 |
| GGT | U/l | 9 - 40 |  | 33 | 33 | 33 | 37 | 28 | 18 | 19 | 18 |
| Total Bilirubin | µmol/l | 7 - 25 |  | 7 | 7 | 8 | 7 | 7 | 5 | 6 | 5 |
| procalcitonine | µg/l | < 0.25 |  |  |  |  |  |  |  | 0.08 |  |
| Quick | % | > 70 |  |  | >100 |  |  |  |  |  |  |
| INR |  |  |  |  | 1 |  |  |  |  |  |  |
| PTT | sec | 26 - 37 |  |  | 27.5 |  |  |  |  | 31.1 |  |
| fibrinogène | g/l | 1.5 - 3.5 |  |  |  |  |  |  |  | 2.4 |  |
| D dimers | ng/ml | 45 - 500 |  |  |  |  |  |  |  | 383 |  |

**Table S5:** Patient 4’s laboratory parameters during hospitalization

| Parameter | Units | Normal range<br>Min/max | D7 | D6 | D5 | D4 | D3 | D2 | D1 |
| --- | --- | --- | --- | --- | --- | --- | --- | --- | --- |
| Erythrocytes | T/l | 4.4 - 6 | 5.31 | 5.51 | 5.45 | 5.5 | 5.45 | 5.3 | 5.54 |
| Hemoglobine | g/l | 140 - 180 | 149 | 155 | 156 | 154 | 156 | 153 | 159 |
| Hematocrite | % | 40 - 52 | 43.4 | 44.8 | 44 | 44.4 | 45.1 | 44.1 | 45.9 |
| MCV | fl | 82 - 98 | 81.7 | 81.3 | 80.7 | 80.7 | 82.8 | 83.2 | 82.9 |
| MCH | pg | 26 - 34 | 28.1 | 28.1 | 28.6 | 28 | 28.6 | 28.9 | 28.7 |
| MCHC | g/l | 320 - 360 | 343 | 346 | 355 | 347 | 346 | 347 | 346 |
| Leucocytes | G/l | 4 - 11 | 3.9 | 3.5 | 3.7 | 4.1 | 4.7 | 5.9 | 6.6 |
| White blood cell count |  |  |  |  |  |  |  |  |  |
| neutrophiles | % | 33 - 80 | 43.6 | 34.7 | 46.9 | 46 | 45 | 61.6 | 80.2 |
| eosinophiles | % | 0 - 5 | 1.3 | 1.4 | 0.5 | 0.2 | 0 | 2 | 0 |
| basophiles | % | 0 - 2 | 0.3 | 0.6 | 0 | 0.2 | 0.2 | 0.3 | 0.3 |
| monocytes | % | 0 - 9 | 9.3 | 11.6 | 12.9 | 12.7 | 14.7 | 13.7 | 12.5 |
| lymphocytes | % | 15 - 60 | 45.5 | 51.7 | 39.7 | 40.9 | 40.1 | 22.4 | 7 |
| neutrophiles-absolute number | G/l | 1.5 - 8 | 1.7 | 1.21 | 1.74 | 1.89 | 2.12 | 3.63 | 5.29 |
| lymphocytes-absolute number | G/l | 1 - 4.5 | 1.77 | 1.81 | 1.47 | 1.68 | 1.88 | 1.32 | 0.46 |
| Thrombocytes | G/l | 150 - 350 | 109 | 113 | 123 | 124 | 113 | 116 | 136 |
| MPV | fl | 8 - 12 | 11.5 | 11.5 | 11.3 | 11.6 | 11.6 | 10.8 | 11.1 |
| C reactive protein | mg/l | 0 - 10 | 1.3 | 1.1 | 1.3 | 1.4 | 2.1 | 5.7 | 7.7 |
| sodium | mmol/l | 136 - 144 | 139 | 137 | 140 | 138 | 138 | 140 | 140 |
| potassium | mmol/l | 3.6 - 4.6 | 3.9 | 3.8 | 4.1 | 3.7 | 3.8 | 3.7 | 3.5 |
| urea | mmol/l | 3.2 - 7.5 | 4.1 | 4.2 | 4.7 | 5.3 | 6 | 6.6 | 6.7 |
| creatinine | μmol/l | 62 - 106 | 94 | 93 | 92 | 94 | 101 | 110 | 95 |
| eGFR (CKD-EPI) | ml/min/1.73m<sup>2</sup> | > 60 | 97 | 99 | 100 | 97 | 89 | 81 | 96 |
| lactate deshydrogénase | U/l | 87 - 210 |  |  |  |  |  |  | 213 |
| AST | U/l | 14 - 50 | 25 | 19 | 22 | 23 | 21 | 23 | 20 |
| ALT | U/l | 12 - 50 | 26 | 25 | 27 | 26 | 26 | 32 | 31 |
| Alcaline phosphatase | U/l | 25 - 102 | 64 | 55 | 53 | 56 | 57 | 58 | 65 |
| GGT | U/l | 9 - 40 | 33 | 34 | 32 | 34 | 35 | 34 | 37 |
| Total Bilirubin | μmol/l | 7 - 25 | 12 | 9 | 10 | 9 | 9 | 7 | 7 |
| procalcitonine | μg/l | < 0.25 |  |  |  |  |  |  |  |
| Quick | % | > 70 |  | >100 |  | 99 | 86 |  | 81 |
| INR |  |  |  | 1 |  | 1 | 1.07 |  | 1.1 |
| PTT | sec | 26 - 37 |  | 25.4 |  | 29 | 27.7 |  | 27.2 |
| fibrinogène | g/l | 1.5 - 3.5 |  |  |  |  | 3.2 |  | 3.3 |

**Table S6:** Patient 5’s laboratory parameters during hospitalization

| Parameter | Units | Normal range | D15 | D13 | D12 | D11 | D10 | D9 | D8 | D6 |
| --- | --- | --- | --- | --- | --- | --- | --- | --- | --- | --- |
|  |  | Min/max |  |  |  |  |  |  |  |  |
| érythrocytes | T/l | 4.4 - 6 | 4.64 | 4.22 | 4.13 |  | 3.92 | 4.05 | 4.2 | 4.73 |
| hémoglobine | g/l | 140 - 180 | 126 | 119 | 118 |  | 113 | 115 | 115 | 132 |
| hématocrite | % | 40 - 52 | 39.2 | 35.6 | 34.9 |  | 33 | 34.5 | 35.7 | 40.1 |
| MCV | fl | 82 - 98 | 84.5 | 84.4 | 84.5 |  | 84.2 | 85.2 | 85 | 84.8 |
| MCH | pg | 26 - 34 | 27.2 | 28.2 | 28.6 |  | 28.8 | 28.4 | 27.4 | 27.9 |
| MCHC | g/l | 320 - 360 | 321 | 334 | 338 |  | 342 | 333 | 322 | 329 |
| Réticulocytes |  |  |  |  |  |  |  |  |  |  |
| réticulocytes | o/oo Ery | 5 - 15 |  |  | 12.6 |  |  |  |  |  |
| réticulocytes-nb.abs | G/l | 20 - 120 |  |  | 52.04 |  |  |  |  |  |
| HFR | % | 0 - 1.5 |  |  | 8.7 |  |  |  |  |  |
| Ret-He | pg | 28 - 35 |  |  | 29.3 |  |  |  |  |  |
| leucocytes | G/l | 4 - 11 | 6 | 5.2 | 5.5 |  | 5.7 | 5.7 | 7 | 12 |
| Répartition leucocytaire |  |  |  |  |  |  |  |  |  |  |
| neutrophiles | % | 33 - 80 |  |  |  |  |  |  |  | 92.6 |
| neutrophiles segmentés | % | 33 - 75 | 64 | 48 | 68 |  | 71 | 75 |  |  |
| neutrophiles non segmnt | % | 0 - 5 | 2 | 2 | 0 |  | 3 | 2 |  |  |
| éosinophiles | % | 0 - 5 | 1 | 9 | 6 |  | 3 | 2 |  | 0 |
| basophiles | % | 0 - 2 | 1 | 0 | 0 |  | 0 | 0 |  | 0.2 |
| monocytes | % | 0 - 9 | 4 | 16 | 16 |  | 12 | 6 |  | 3.4 |
| lymphocytes | % | 15 - 60 | 25 | 20 | 9 |  | 11 | 15 |  | 3.8 |
| myélocytes | % | 0 - 0 | 3 | 5 | 1 |  |  |  |  |  |
| cellules réparties |  |  | 100 | 100 | 100 |  | 100 | 100 |  |  |
| neutrophiles-nb.abs | G/l | 1.5 - 8 |  |  |  |  |  |  |  | 11.11 |
| neutro segmentes nb.abs | G/l | 1.5 - 7.5 | 3.84 | 2.5 | 3.74 |  | 4.05 | 4.28 |  |  |
| neutro non seg.-nb.abs | G/l | 0 - 0.5 | 0.12 | 0.1 | 0 |  | 0.17 | 0.11 |  |  |
| lymphocytes-nb.abs | G/l | 1 - 4.5 | 1.5 | 1.04 | 0.5 |  | 0.63 | 0.86 |  | 0.46 |
| Thrombocytes |  |  |  |  |  |  |  |  |  |  |
| thrombocytes | G/l | 150 - 350 | 489 | 419 | 352 |  | 284 | 262 | 235 | 222 |
| MPV | fl | 8 - 12 |  | 9 | 9.2 | 9.3 | 9.3 | 9.5 | 9.4 | 9.5 |
| C reactive protein | mg/l | 0 - 10 |  | 26.7 | 44 | 70.1 | 76.4 | 89.2 | 144.4 | 180.5 |
| sodium | mmol/l | 136 - 144 |  | 138 | 142 | 141 | 139 | 141 | 140 | 138 |
| potassium | mmol/l | 3.6 - 4.6 |  | 3.9 | 4 | 3.6 | 3.4 | 3.7 | 3.9 | 3.8 |
| urea | mmol/l | 3.2 - 7.5 |  | 3 | 3.2 | 2.8 | 2.5 | 3.3 | 4.5 | 4.3 |
| creatinine | μmol/l | 62 - 106 |  | 58 | 55 | 59 | 55 | 58 | 59 | 63 |
| eGFR (CKD-EPI) | ml/min/1.73m<sup>2</sup> | > 60 |  | 101 | 103 | 100 | 103 | 101 | 100 | 98 |
| Troponine T ultra sensible | ng/l | < 14 |  |  |  | 5 [E] |  |  |  |  |
| lactate deshydrogénase | U/l | 87 - 210 |  |  |  |  | 191 |  |  |  |
| AST | U/l | 14 - 50 |  | 44 | 39 | 33 |  | 30 | 26 | 24 |
| ALT | U/l | 12 - 50 |  | 45 | 37 | 29 |  | 23 | 16 | 16 |
| Alcaline phosphatase | U/l | 25 - 102 |  | 58 | 59 | 68 |  | 69 | 58 | 49 |
| GGT | U/l | 9 - 40 |  | 61 | 64 | 75 |  | 68 | 52 | 38 |
| Total Bilirubin | μmol/l | 7 - 25 |  | 5 | 6 | 18 |  | 9 | 8 | 9 |
| procalcitonine | μg/l | < 0.25 |  |  |  |  |  | 0.19 |  |  |
| Quick | % | > 70 |  |  |  |  |  | >100 |  | >100 |
| INR |  |  |  |  |  |  |  | 1 |  | 1 |
| PTT | sec | 26 - 37 |  |  |  |  |  | 25.2 |  | 26.8 |
| fibrinogène | g/l | 1.5 - 3.5 |  |  |  |  |  |  |  | 8.4 |

**Figure S1:**
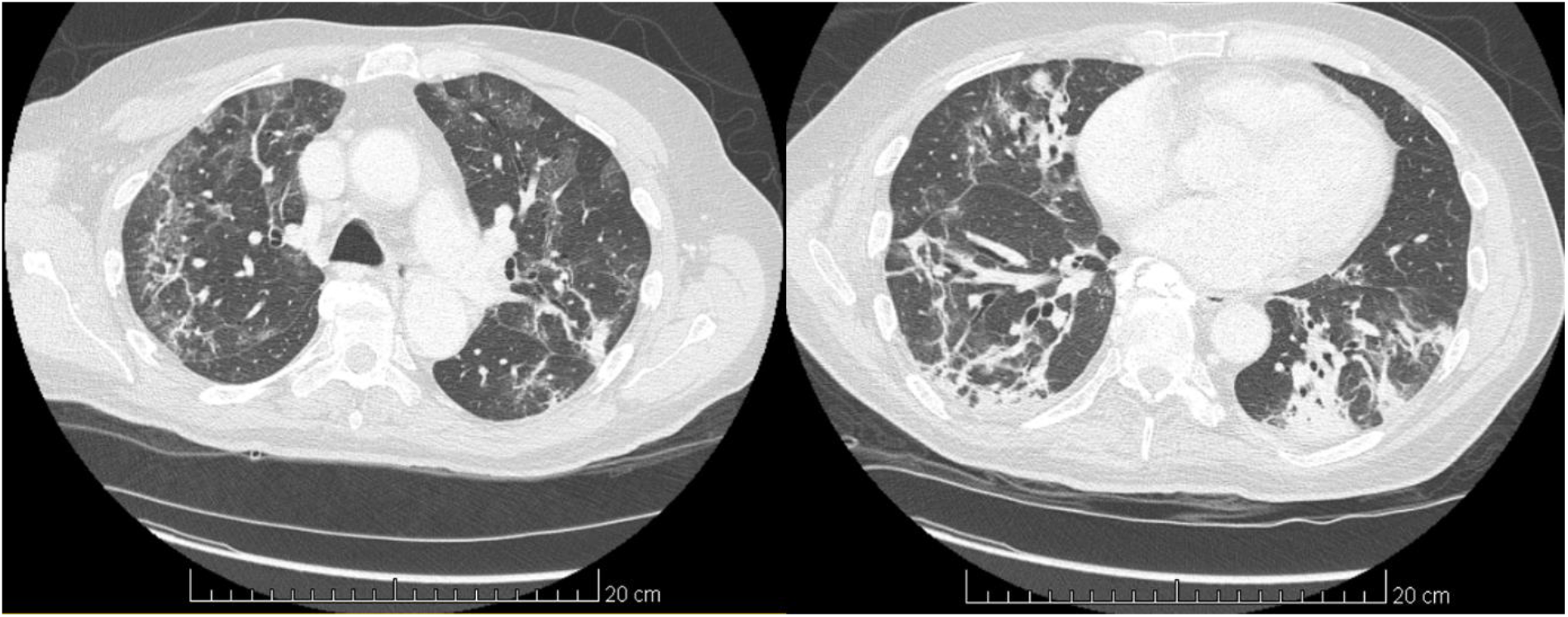
CT-scan of patient 5 showing (A) bilateral patchy ground-glass opacities, mainly in the upper lobes, and (B) postero-basal consolidation, more marked on the left side.

**Figure S2:**
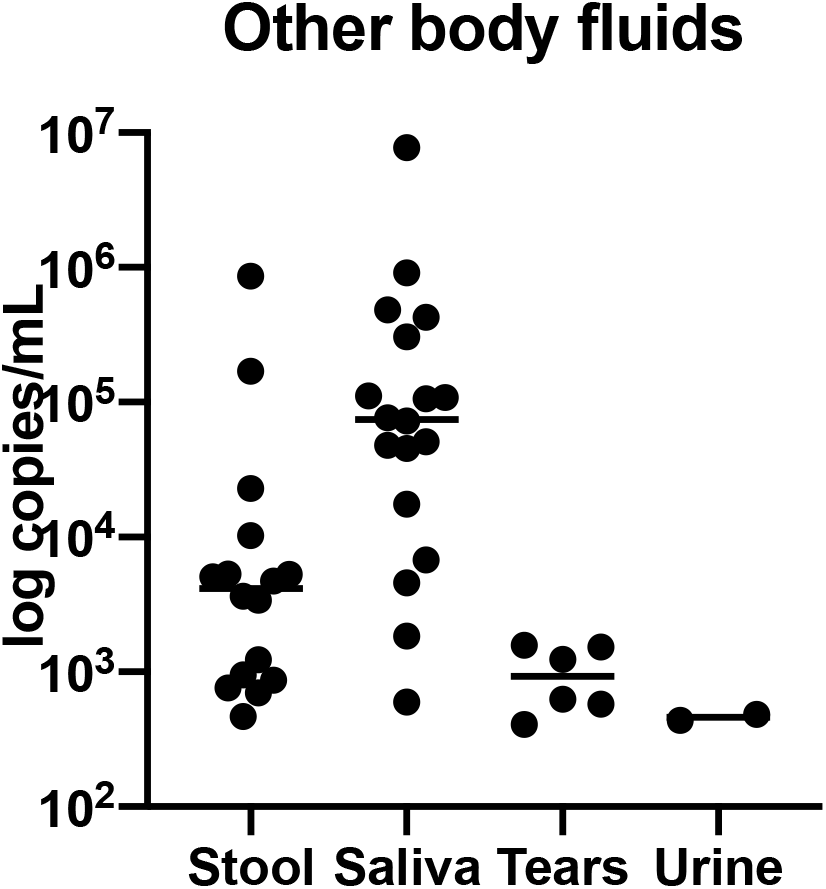
viral load in saliva, stool, tears and urine

**Figure S3:**
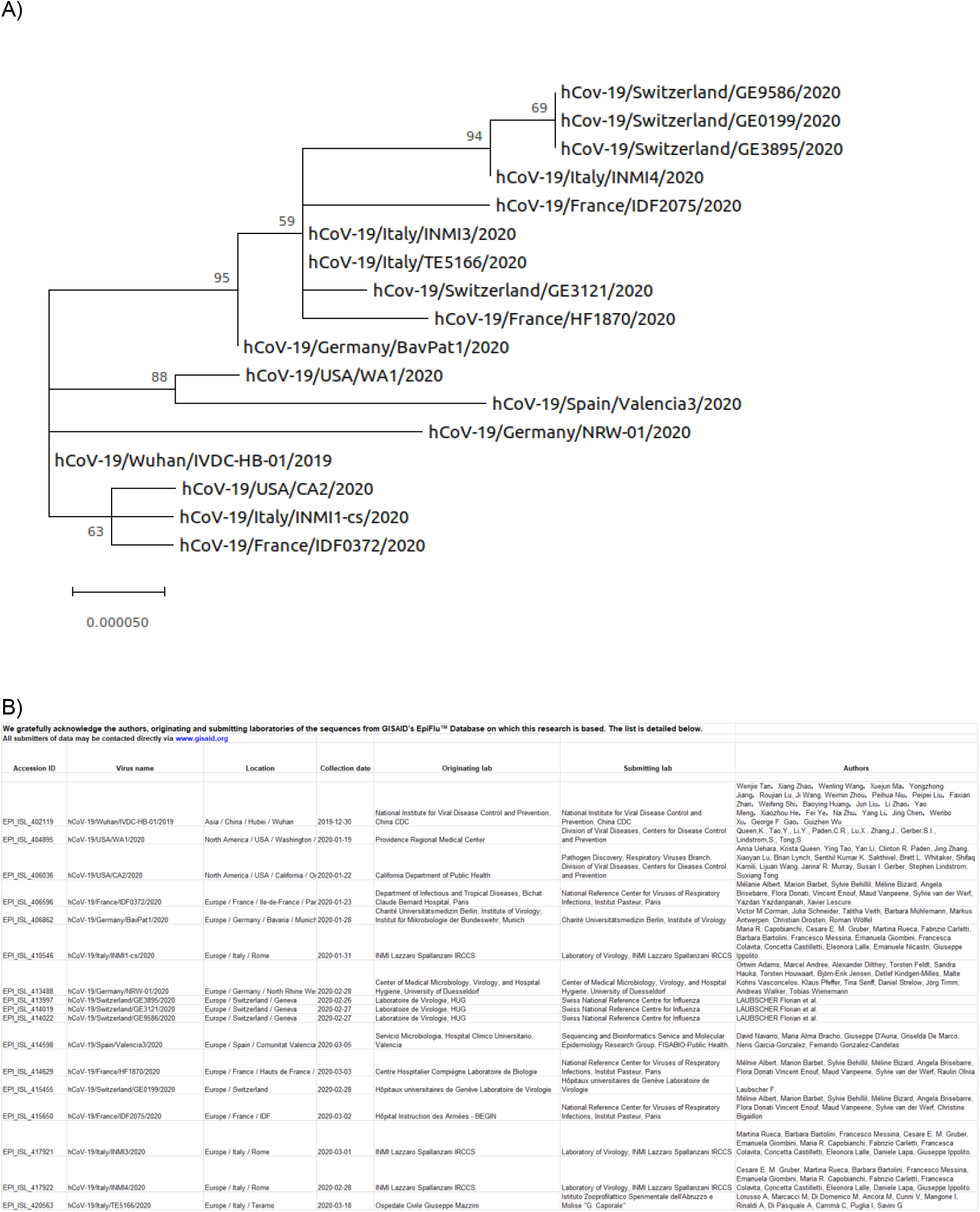
Phylogenetic tree constructed on the basis of SARS-CoV-2 complete genome sequences. A) The Evolutionary analyses were conducted in MEGA X using the Maximum Likelihood method and Hasegawa-Kishino-Yano model. All sequences were downloaded from GISAID database (for more information refer to the gisaid_hcov-19_acknowledgement_table below). Scale bar indicates nucleotide substitutions per site. B) GISAID HcoV-19 acknowledgement table

